# Outcomes of a Smartphone-based Application with Live Health-Coaching Post-Percutaneous Coronary Intervention

**DOI:** 10.1101/2020.11.08.20217653

**Authors:** Kaavya Paruchuri, Phoebe Finneran, Nicholas A Marston, Emma W Healy, John Andreo, Ryan Lynch, Alexander J Blood, Maeve Jones-O’Connor, Bradley Lander, Noreen Kelly, Maria T. Vivaldi, Kate Traynor, Stephen Wiviott, Pradeep Natarajan

## Abstract

**Background:** The interval between inpatient hospitalization for symptomatic coronary artery disease (CAD) and post-discharge office consultation is a vulnerable period for adverse events.

**Methods:** Content was customized on a smartphone app-based platform for hospitalized patients receiving PCI which included education, tracking, reminders and live health coaches. We conducted a single-arm open-label pilot study of the app at two academic medical centers in a single health system, with subjects enrolled 02/2018-05/2019 and 1:3 propensity-matched historical controls from 01/2015-12/2017. To evaluate feasibility and efficacy, we assessed 30-day hospital readmission (primary), outpatient cardiovascular follow-up, and cardiac rehabilitation (CR) enrollment as recorded in the health system. Outcomes were assessed by Cox Proportional Hazards model.

**Findings:** 118 of 324 eligible (36·4%) 21-85 year-old patients who underwent PCI for symptomatic CAD who owned a smartphone or tablet enrolled. Mean age was 62.5 (9·7) years, 87 (73·7%) were male, 40 of 118 (33·9%) had type 2 diabetes mellitus, 68 (57·6%) enrolled underwent PCI for MI and 59 (50·0%) had previously known CAD; demographics were similar among matched historical controls. No significant difference existed in all-cause readmission within 30 days (8·5% app vs 9·6% control, ARR -1.1% absolute difference, 95% CI -7·1-4·8, p=0·699) or 90 days (16·1% app vs 19·5% control, p=0.394). Rates of both 90-day CR enrollment (HR 1·99, 95% CI 1·30-3·06) and 1-month cardiovascular follow up (HR 1·83, 95% CI 1·43-2·34) were greater with the app. Weekly engagement at 30- and 90-days, as measured by percentage of weeks with at least one day of completion of tasks, was mean (SD) 73·5% (33·9%) and 63·5% (40·3%). Spearman correlation analyses indicated similar engagement across age, sex, and cardiovascular risk factors.

**Interpretations:** A post-PCI smartphone app with live health coaches yielded similarly high engagement across demographics and safely increased attendance in cardiac rehabilitation. Larger prospective randomized controlled trials are necessary to test whether this app improves cardiovascular outcomes following PCI.

**Funding:** National Institutes of Health, Boston Scientific.

**Clinical Trial Registration:** NCT03416920 (https://clinicaltrials.gov/ct2/show/NCT03416920).

## INTRODUCTION

Myocardial infarction (MI) is a leading cause of morbidity and mortality in the United States and worldwide.^1,2^ Acute management of MI comprises pharmacological and revascularization strategies to reduce recurrent ischemic events, including cardiovascular death.^3,4^ Secondary prevention measures include adherence to guideline-based pharmacologic strategies, management of cardiovascular risk factors, diet and lifestyle counseling, and participation in cardiac rehabilitation. Both patient- and treatment-specific factors are associated with short-term adverse events.

Cardiac rehabilitation (CR) has been demonstrated to reduce both mortality rates and hospital admissions after coronary artery disease (CAD) events, such as MI, by providing a comprehensive secondary prevention framework in randomized controlled trials.^5–7^ Despite the known benefits, barriers to CR remain, and are highlighted by low referral, enrollment and completion rates.^8,9^ Non-commercial insurance status, non-white race, older age, female gender, distance to facility, medical comorbidities, and geographic socioeconomic factors are correlated with lower CR utilization rates.^10,11^ Among level I guideline recommendations, CR consistently attains the lowest adherence rates.^12^

Mobile health platforms have been previously proposed as novel care strategies with recent rapid adoption to minimize SARS-CoV-2 transmission.^13^ Smartphone and internet-based programs may safely deliver elements of CR.^14,15^ Virtual strategies may improve access to care and facilitate lifestyle modification.^16^ Smartphone apps concurrent with CR may improve CR adherence.^17^ Randomized controlled trials to evaluate the potential synergy of digital health platforms with traditional CR are underway.^18–20^ The Million Hearts Cardiac Rehabilitation Collaborative aims to improve CR enrollment itself, but whether engagement earlier through digital health strategies may support this goal is not presently known.^21^

Here, we customized an app to address the inpatient-outpatient transition post-PCI to improve enrollment of CR. Our goals were to assess feasibility of smartphone application (app) deployment during inpatient hospitalization and evaluate efficacy of the app in improving CR enrollment as well as short-term safety of engagement with third-party health coaches compared to matched historical controls.

## METHODS

### Study Participants

The present study is an open-label, single-arm, multi-center clinical trial with stratified enrollment and matched historical controls (NCT03416920). The study protocol was approved by the Massachusetts General Hospital Institutional Review Board (#2017P002582).

Between 02/2018 and 05/2019, patients were identified via electronic health records on the day after PCI for either non-MI symptomatic CAD or acute MI at study sites (Massachusetts General Hospital [MGH] and Brigham and Women’s Hospital [BWH]) within the Mass General Brigham (MGB, formerly Partners) healthcare system. Enrollment for symptomatic CAD, in the absence of MI, was capped at 50 participants per prespecified protocol. English-speaking patients aged 21-70 years who owned a smartphone or tablet and had a longitudinal MGB primary care provider were considered for inclusion. Inclusion criteria were later liberalized to include patients aged up to 85 years and without MGB primary care providers due to patient interest. Patients with recent (within 1 month) use of illicit substances or alcohol abuse, in-hospital acute MI, known pregnancy, dementia or cognitive delay or incarceration were excluded.

Historical controls were ascertained by query of a centralized clinical data repository to identify a historical patient population who met study eligibility criteria, except smartphone or tablet possession information was not available, who underwent PCI from 01/01/2015 through 12/31/2017 at the two study sites. Historical controls were then identified in a 1:3 manner via propensity matching^22^ (nearest neighbor method, MatchIt Package v2·0·2, RStudio) on key demographic criteria including age, sex, state of residence (Massachusetts [MA]/non-MA), PCI Type (MI/symptomatic CAD), diabetes mellitus type 2 (DM2), MGB primary care provider (yes/no), insurance type (commercial/Medicaid/Medicare/other) and PCI site (MGH/BWH). Identified controls were then validated for all demographics by physician manual chart review; 1 identified control was removed due to validation failure. Balance diagnostics were verified using standardized mean difference (cobalt package, v4·2·2, RStudio).^23^

### Digital Health Platform

Content on a digital health platform from a Massachusetts-based health care technology company (Wellframe, Boston, MA) was customized for hospitalized patients receiving percutaneous coronary intervention (PCI).^24^ The mobile health platform consisted of a patient-facing mobile app, a clinical dashboard and a suite of clinical programs with configurable rules (**Supplementary Figure 1**). The patient mobile app featured a personalized adaptive daily health checklist that included reminders to engage in health behaviors and a series of personalized, interactive surveys, articles and encouragement (**Supplementary Text**). The app was developed using user-experience and design principles to ensure operability by a wide variety of users.

### Trial Procedures

Enrolled participants in the intervention arm provided written and verbal consent, and the app was installed on their personal smartphone or tablet prior to hospital discharge. Patients had active accounts on the study app for 90 days, after which each participant was contacted once by a member of the study staff. During this phone call, staff asked a series of standardized questions to determine clinical outcomes.

Manual chart review by study physicians was conducted for all participants in both the interventional and historical control arms to harmonize outcome adjudication between both groups. In both groups, hospitalization within an MGB hospital with documentation of date within 30- or 90-days was used for readmission rates.

Emergency department (ED) or urgent care (UC) visit with subsequent hospitalization was counted as a single readmission. Admission or presentation with documentation of a chief complaint consistent with cardiac etiology (i.e. CAD, CHF, arrhythmia), admission requiring cardiac consultation or admission to a cardiology ward unit was coded as a cardiovascular hospitalization.

### Trial Outcomes

We primarily assessed the safety of the app, as assessed by 30-day all-cause hospital readmission, and secondarily 90-day all-cause hospital readmission. We also secondarily assessed enrollment in CR at an MGB site within 90 days and follow up with an MGB cardiologist within 1 month of hospitalization (to harmonize outcome ascertainment between the intervention group and the historical control group). We evaluated feasibility metrics including conversion rate, or the study enrollment rate among the total number of eligible patients. Among the participants in the intervention group, we evaluated various engagement metrics **(Supplementary Table 1)**.

### Statistical Analyses

Differences in baseline characteristics between study groups were first assessed by Fisher’s exact tests or two-sample t-tests (if variables were continuous). Clinical outcome risks, including 30- and 90-day all cause and CV readmission, 90-day CR enrollment, and 1-month outpatient cardiology follow-up were estimated between study groups by Cox proportional hazards models. Models were adjusted for age, sex, insurance type, state of residence, DM2, and primary care provider practice location. Baseline was set at date of PCI, and if the aforementioned events did not occur, last follow-up was established at date of death or last follow-up within 90 days (data were right censored at 90 days if events did not occur). The proportional hazards assumption was evaluated by examining Schoenfeld residuals and was met.

Spearman correlation analysis was used to correlate engagement metrics with age, sex, reason for PCI (MI, non-MI symptomatic CAD) or known cardiovascular risk factors (hypertension, hyperlipidemia, DM2, peripheral arterial disease and known CAD). Mean daily engagement was then dichotomized into top 25^th^ percentile and bottom 75^th^ percentile. Multivariate ANOVA was used to correlate quartiles of engagement with 90-day CR enrollment, 90-day all cause readmission and 1-month outpatient follow up.

Statistical significance was assigned at two-sided alpha=0·05. Analyses were performed in RStudio (v1·2·5001, RStudio Inc, Boston MA; R version 3·6·1, R Foundation, Vienna, Austria).

## RESULTS

### Baseline Characteristics

1,994 patients were screened at time of PCI at two study sites. 1,676 patients were excluded primarily due to lack of consent (650), elective PCI after cap attained (370), advanced age (209), or deemed not suitable by the treating team (208) (**Supplementary Figure 2**). The remaining 324 were eligible and approached, 118 of whom (36·4%) were enrolled in the study. 68 of 118 (57·6%) underwent PCI for MI, and 29 of 118 (24·6%) were for ST elevation MI. Mean (standard deviation [SD]) age was 62.5 (9·7) years, 87 (73·7%) were male, 40 (33·9%) had DM2, and 59 (50·0%) had previously known CAD. Demographics were similar between enrolled participants and historical controls (**Table 1**). Adjusted standard mean differences for covariates were < 0·25 (**Supplementary Figure 3**).

**Table 1:** Baseline characteristics of all subjects. Demographics across both historical controls and participants enrolled in the study app were similar. Fisher’s exact testing was used to evaluate differences across groups (†Two-sample t-testing was used for continuous variables). (SD = Standard Deviation, Min = Minimum, Max = Maximum, MI = PCI for Acute Myocardial Infarction, Elective = PCI for Symptomatic Coronary Artery Disease without Acute Myocardial Infarction, MGH = Massachusetts General Hospital, BWH = Brigham and Women’s Hospital, MA = Massachusetts)

|  | Historical<br>Control Group<br>(n=343) | Intervention<br>Group<br>(n=118) | p-value<br>(Fisher's<br>exact test) |
| --- | --- | --- | --- |
| **Age, years** |  |  |  |
| Mean (SD) | 63.4 (10.3) | 62.5 (9.72) | 0.396† |
| Median<br>[Min, Max] | 64.0<br>[35.0, 84.0] | 63.5<br>[38.0, 80.0] |  |
| **Sex** |  |  |  |
| Male | 247 (72.0%) | 87 (73.7%) | 0.811 |
| Female | 96 (28.0%) | 31 (26.3%) |  |
| **PCI Type** |  |  |  |
| MI | 217 (63.3%) | 68 (57.6%) | 0.322 |
| Elective | 126 (36.7%) | 50 (42.4%) |  |
| **Site** |  |  |  |
| MGH | 267 (77.8%) | 93 (78.8%) | 0.898 |
| BWH | 76 (22.2%) | 25 (21.2%) |  |
| **Ethnicity** |  |  |  |
| White | 273 (79.6%) | 102 (86.4%) | 0.649 |
| Black | 14 (4.1%) | 3 (2.5%) |  |
| Hispanic | 11 (3.2%) | 1 (0.8%) |  |
| Asian | 10 (2.9%) | 3 (2.5%) |  |
| Other | 19 (5.5%) | 6 (5.1%) |  |
| Unknown | 16 (4.7%) | 3 (2.5%) |  |
| **Insurance Type** |  |  |  |
| Commercial | 204 (59.5%) | 72 (61.0%) | 0.995 |
| Medicaid | 24 (7.0%) | 8 (6.8%) |  |
| Medicare | 110 (32.1%) | 37 (31.4%) |  |
| Other | 5 (1.5%) | 1 (0.8%) |  |
| **State** |  |  |  |
| MA | 279 (81.3%) | 99 (83.9%) | 0.581 |
| Non-MA | 64 (18.7%) | 19 (16.1%) |  |
| **MGB Primary Care Provider** |  |  |  |
| Yes | 160 (46.6%) | 54 (45.8%) | 0.915 |
| No | 183 (53.4%) | 64 (54.2%) |  |
| **Diabetes Mellitus Type 2** |  |  |  |
| Yes | 128 (37.3%) | 40 (33.9%) | 0.579 |
| No | 215 (62.7%) | 78 (66.1%) |  |
| **Hypertension** |  |  |  |
| Yes | 244 (71.1%) | 81 (68.6%) | 0.640 |
| No | 99 (28.9%) | 37 (31.4%) |  |
| **Hyperlipidemia** |  |  |  |
| Yes | 266 (77.6%) | 88 (74.6%) | 0.528 |
| No | 77 (22.4%) | 30 (25.4%) |  |
| **Peripheral Arterial Disease** |  |  |  |
| Yes | 31 (9.0%) | 7 (5.9%) | 0.337 |
| No | 312 (91.0%) | 111 (94.1%) |  |
| **Known Coronary Artery Disease** |  |  |  |
| Yes | 156 (45.5%) | 59 (50.0%) | 0.649 |
| No | 187 (54.5%) | 59 (50.0%) |  |

### Study App Engagement

Engagement with the app, as defined by completion of at least one task either daily or weekly, fell in a parabolic distribution (**Supplementary Figure 4**). Patients set up a mean (SD) of 3·45 (6·23) medication reminders per day. Patients sent a median [IQR] of 1 [9] message and the clinical care team sent a median [IQR] of 12 (11) messages per participant. The number of messages sent by health coaches was highly correlated with the number of messages sent by individual participants (Spearman r^2^ =0·82, p<2·2×10^−16^, **Supplementary Figure 5**). Spearman correlation analyses revealed no significant associations between number of messages per participant and age, sex or known cardiovascular risk factors or PCI indication.

Engagement rates were durable across 30- and 90-day periods (**Table 2**). 30- and 90-day content completion rates, as measured by percentage of articles opened by participants, were mean (SD) 57·8% (36·5%) and 52·9% (36·1%), respectively. Medication adherence rates at 30- and 90-days, as measured by percentage of medication reminder tasks completed by participants, were mean (SD) 76·3% (31·3%) and 69·9% (30·7%) respectively. Survey completion rates at 30- and 90-days, as measured by percentage of survey tasks completed, were mean (SD) 52·4% (39·8%) and 46·0% (37·9%). The percentage of days participants met their physical activity goal was mean (SD) 53·3% (41·4%) at 30 days and 52·3% (40·3%) at 90 days. Daily engagement rates at 30- and 90-days, as measured by percentage of days with at least one task completion, were mean (SD) 55·4% (40·5%) and 48·2% (40·6%). Weekly engagement at 30- and 90-days, as measured by percentage of weeks with at least one day of completion of tasks, was mean (SD) 73·5% (33·9%) and 63·5% (40·3%) respectively (**Figure 1)**. Spearman correlation analyses revealed no significant correlations between engagement metrics and age, sex or known cardiovascular risk factors (**Supplementary Figure 6**).

**Table 2:** Engagement with app. Engagement metrics were similar over the entire study period suggesting stable usage of the app amongst users who were engaged early. (SD = Standard Deviation)

| **ENGAGEMENT METRIC** | **0-30 DAYS**<br>**MEAN (SD)** | **0-90 DAYS**<br>**MEAN (SD)** |
| --- | --- | --- |
| Content Completion<br>*(Percentage of articles opened)* | 57.8% (36.5%) | 52.9% (36.1%) |
| Medication Adherence<br>*(Percentage of medication reminder tasks completed)* | 76.3% (31.3%) | 69.9% (30.7%) |
| Survey Completion<br>*(Percentage of survey tasks completed)* | 52.4% (39.8%) | 46.0% (37.9%) |
| Physical Activity Completion<br>*(Percentage of self-reported days physical activity goal was met)* | 53.3% (41.4%) | 52.3% (40.3%) |
| Daily Engagement<br>*(Percentage of days with completion of at least one task)* | 55.4% (40.5%) | 48.2% (40.6%) |
| Weekly Engagement<br>*(Percentage of weeks with at least one day of completion of one task)* | 73.5% (33.9%) | 63.5% (40.3%) |

**Figure 1:**
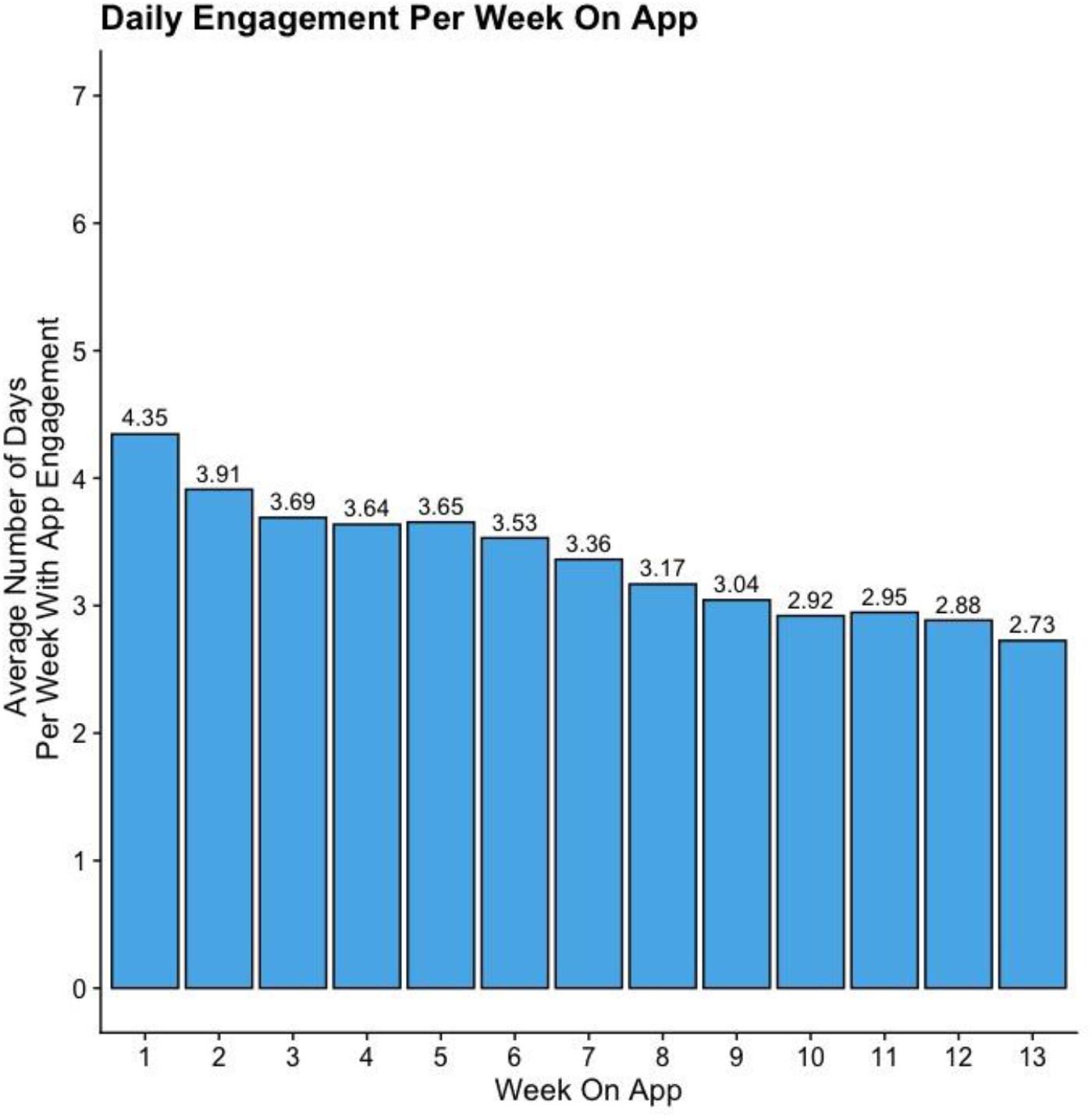
Weekly app engagement. Patients’ interaction with the app decreased gradually over time but the overall rate was stable amongst patients who engaged early. Engagement peaked at 4.35 days in the first week and gradually decreased to 2.73 days by the last week of the study period. (Engagement = a day with completion of at least one task; Week = 7-day period starting from enrollment).

### Study Intervention Outcomes

There was an increased rate in 90-day enrollment in CR at an MGB site (29·7% vs 16·3%, p=1·5×10^−3^) (**Figure 2A**) for the intervention group versus historical controls. Among those who enrolled in CR, we observed that 21/35 (60·0%) in the intervention group and 23/56 (41·1%) in the historical control group completed CR (p=0·09). We observed consistent relationships within both sites **(Supplementary Figure 7)**. Among those who enrolled in CR, demographics were largely similar between the intervention and historical control groups (**Supplementary Table 2**).

**Figure 2:**
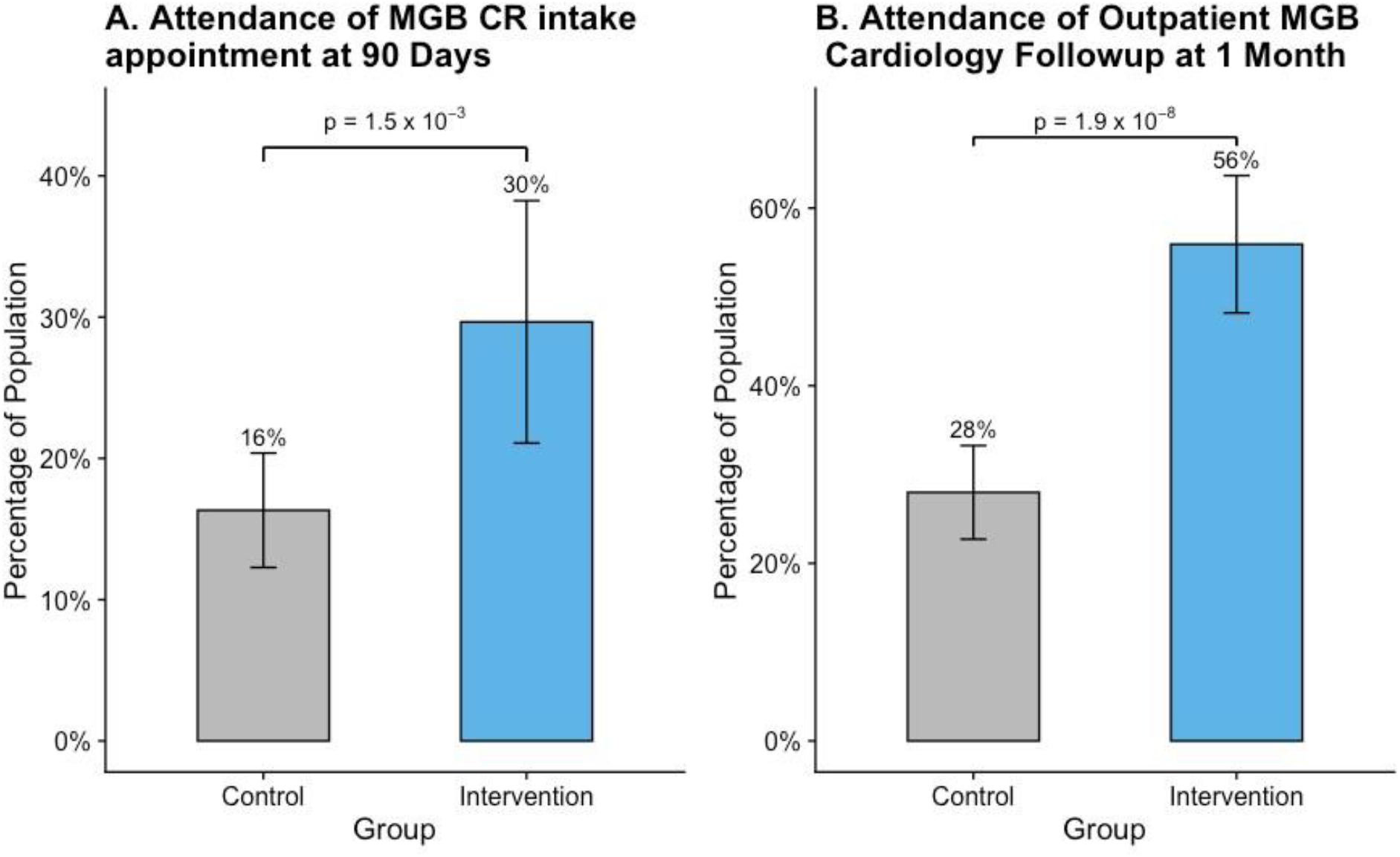
A smartphone app and longitudinal cardiovascular care. (A) Two-fold increase in attendance of cardiac rehabilitation intake and (B) two-fold increase in 1-month outpatient cardiovascular follow up in the intervention group. Error bars represent confidence intervals. (MGB = Mass General Brigham)

For the intervention arm, phone follow-up was also performed and successful for 75 (63·5%) participants. Of those in the intervention group who were successfully contacted by phone, 46 (61·3%) enrolled in CR at any site within 90 days. In this group, 22 (29·3%) specifically participated in CR at an MGB site.

66 (55·9%) participants in the intervention group attended 1-month outpatient MGB cardiology follow-up compared to 96 (28·0%) among historical controls (p=1·9×10^−8^) (**Figure 2B**). For those who attended a 1-month outpatient MGB cardiology follow-up, the mean number of days from PCI date to first outpatient MGB cardiology follow-up was mean (SD) 18·1 (7·43) in the intervention group and 19·1 (7·61) in the historical control group (p=0·39). Of those in the intervention group who were successfully contacted by phone, 73 of 75 (97·3%) followed up with a cardiologist at any site. In this group, 58 (77·3%) followed up at an MGB cardiology site.

We observed no significant difference in all-cause readmission rates within 30 or 90 days at an MGB hospital between the historical control and intervention groups (33 [9·6%] vs 10 [8·5%], p=0·699; 67 [19·5%] vs 19 [16·1%], p=0·394) (**Figure 3A**). We also observed no significant difference in 30- or 90-day cardiovascular-related readmission rates between the historical control and intervention groups (28 [8·2%] vs 10 [8·5%], p=0·930; 46 [13·4%] vs 18 [15·3%], p=0·191 respectively) (**Figure 3B**). In the intervention arm, daily engagement rate did not predict 90-day all cause readmission (p=0·289), 90-day CR enrollment (p=0·969) nor 1-month outpatient cardiology follow-up (p=0·803) at MGB facilities (**Supplementary Figure 8**).

**Figure 3:**
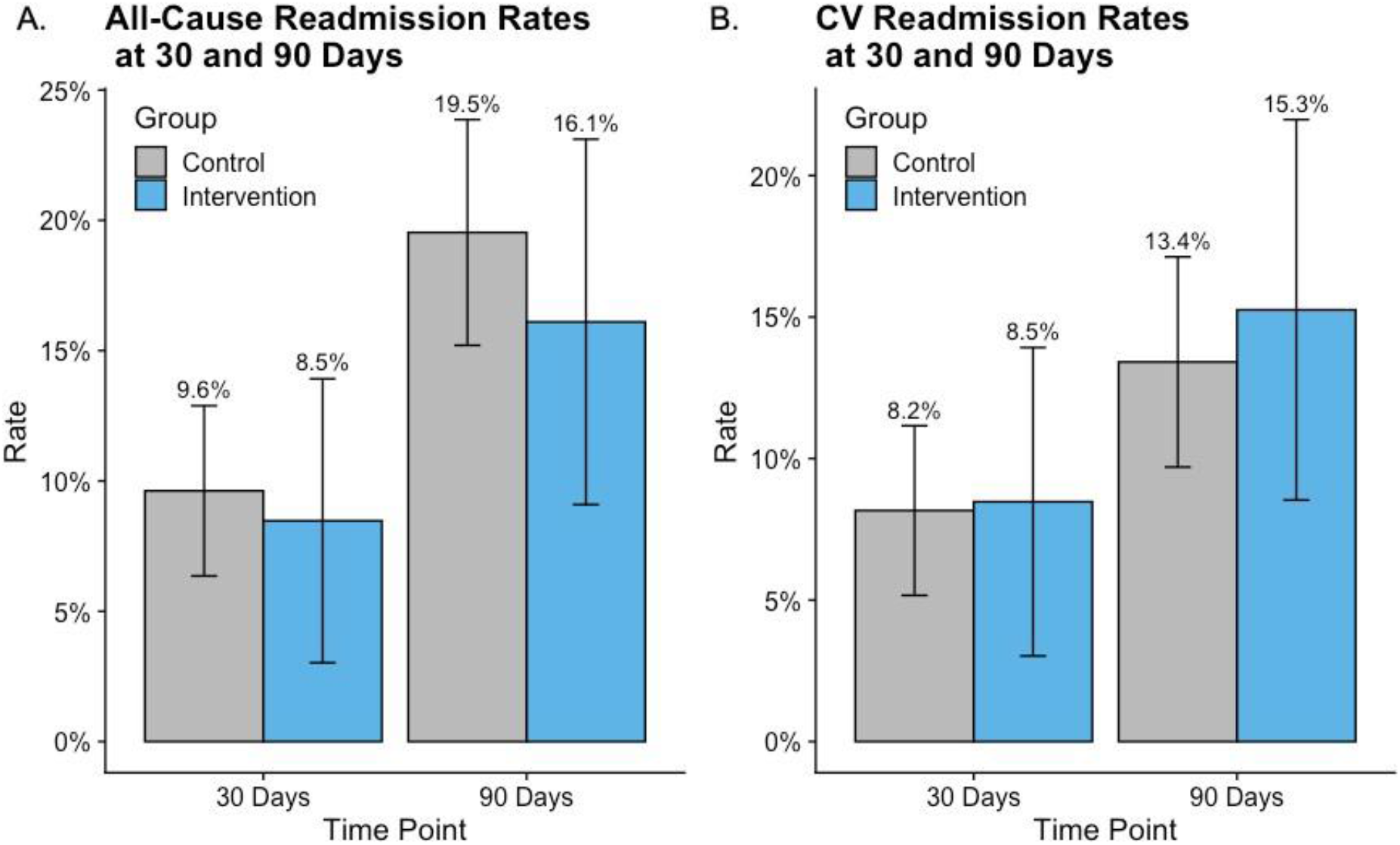
Use of a smartphone app and readmission rates post-PCI. No significant difference in (A) all-cause readmissions or (B) cardiovascular readmission to MGB facilities at either 30- or 90-days post-PCI. Error bars represent confidence interval.

## DISCUSSION

The present smartphone app including third-party health coaches helped bridge a care gap shortly after hospital discharge for PCI and facilitated participation in CR. 1 in 3 individuals approached accepted the app and 52% of individuals surveyed were still using the app daily at 90 days. For every 8 individuals who used the app, 1 additional individual enrolled in CR compared to historical controls. Importantly, use of an app with third-party health coaches did not lead to excess short-term readmissions in the studied population.

Our results may permit several conclusions regarding the prevention of cardiovascular disease using digital health strategies. First, onboarding patients for digital health care solutions during inpatient hospitalization is feasible. Prior digital health initiatives have largely targeted patients in the ambulatory setting or at time of enrollment at CR with the aim of promoting preventive activities and promoting long-term maintenance.^18^ However, the weeks to months between hospital discharge and outpatient CR or cardiovascular follow-up after the index event represents a missed opportunity.^25^ Our study indicates an opportunity to initiate comprehensive cardiovascular disease prevention earlier and facilitate CR enrollment.^9,19^ Use of EHR-integrated digital health tools to engage patients and clinicians in discharge planning tasks has been shown to be feasible, acceptable and valuable.^26^ Therefore, creating standardized clinical workflows to incorporate onboarding with patient-facing digital health tools at time of discharge may enhance usability of patient discharge paperwork and potentially clinical outcomes.

Second, our study shows that patients are willing to engage both with an app targeting cardiovascular disease as well as third-party health coaches. Previous digital health interventions involving automated one-way texting/messaging and structured questionnaires have had mixed success in promoting durable lifestyle changes.^27^ Although behavioral modification studies and patient opinions have suggested that personalized virtual feedback is felt to be the most effective means for enacting change, very few studies have been able to create individualized interventions as they require human guidance.^28,29^ The use of physicians or physician extenders in the role of virtual health coaches within prior studies of digital health interventions in cardiology has allowed for personalization at the cost of limited scalability.^30^ Our study is novel in its utilization of third-party health coaches external to the health system to support patient engagement with minimal burden to clinical staff. Engagement with our framework was markedly greater compared to population-based preventive cardiovascular interventions to-date using apps. In a study of nearly 50,000 individuals in the community, surveys and study procedures over 7 days were only completed by 3-10% of participants.^31,32^ Apps aligned with the healthcare teams that provide passive and active engagement designed for patients with cardiovascular disease may provide more durable effects.

Third, patient engagement with digital health tools may not be predicted by traditional cardiovascular risk factors including comorbidities. There was notably no difference in engagement with the app by age despite preconceived notions of technologic proficiency in older patients. There are few reports of older adults (age >65 years) reporting difficulty with digital health technology but this may reflect their lack of inclusion in clinical studies.^33,34^ Further study is needed to determine whether newer mobile technologies can be adopted into populations previously not represented, such as the elderly and medically complex, who have historic disparities in CR enrollment.^11^

Fourth, despite the resource-rich environment of the app, there was still increased patient engagement with healthcare teams as measured by outpatient follow-up rate and increased adherence to guideline-supported care plans, such as CR enrollment, compared with historic controls. There was no evidence of excess cardiovascular risk, as measured by short-term readmissions, suggesting that participants did not use the app as a surrogate for care. Whether this ‘bridging’ strategy may optimize long-term risk reduction through facilitating CR enrollment, cardiovascular follow-up, patient education, and motivational coaching requires longer term study in randomized controlled trials.

While our study has several strengths, it also has limitations. First, participants who opt to enroll in this novel study may be more engaged in health maintenance compared to those who decline. Second, reliance on historical controls with propensity matching may lead to imbalance of unavailable phenotypes or systematic healthcare changes. We focused on relatively short duration, contemporary time periods at two sites, and matched used several clinical features related to health-related behaviors.^10^ Persistently low rates of readmission for the two groups indicate the lack of major new systematic endeavors in addressing readmission rates between the two periods. Additionally, there were no other formal systemic endeavors targeting CR enrollment rates within the healthcare system studied. Third, use of historical controls permitted us to only ascertain clinical events from the electronic health record and thus, from within our health system. Power is maximized since the studied health system is the largest in New England, but this may have led to reduced absolute rates observed. If similar trends were observed outside of the healthcare system, then greater absolute risk differences may be truly present with the current data thus biasing our results to the null. Finally, given the small sample size and short-term follow up, we cannot exclude the possibility of reduced power in the assessment of clinical cardiovascular outcomes.

In conclusion, a post-PCI smartphone app, with live health coaches, deployed upon discharge yields similarly high engagement across demographics. Compared to historical controls, use of the app did not reduce short-term hospital readmission but was associated with two-fold higher attendance in CR. Prospective randomized controlled trials are necessary to test whether this digital health platform improves cardiovascular outcomes following PCI.

## Data Availability

The data dictionary will be available in the supplementary material. Individual level data will not be made available due to privacy concerns.

## Data Access, Responsibility and Analysis

P.N and K.P. had full access to all the data in the study and take responsibility for the integrity of the data and the accuracy of the data analysis.

## Data Sharing

A data dictionary defining types of data collected during this study will be made available to others in supplementary text with publication. Individual level participant data will not be made available to others due to privacy concerns. Study protocol and statistical analysis plan will be made available to any academic researchers who request it from corresponding author.

## Research in context

### Evidence before this study

Although mobile health platforms have been proposed as novel health care strategies, evidence to indicate feasibility and safety in transitional care is limited. Smartphone apps with third-party health coaches to improve the inpatient to outpatient care transition for cardiovascular disease within existing clinical workflows have not been well described. Whether such apps can bridge high risk care gaps and improve adoption of guideline-supported therapies is unclear.

### Added value of this study

This study reports the results of a pilot study utilizing a smartphone app with live health coaching provided at hospital discharge in 118 patients post-percutaneous coronary intervention. The app delivers customized education content, allows medication and step tracking, and provides personalized feedback from non-clinician live health coaches. This study shows that an app improves hospital discharge with a high degree of engagement and improved transition to outpatient care and therapies, such as increased cardiac rehabilitation enrollment, compared with historical controls. There was no evidence of excess cardiovascular risk with use of the app, as measured by short-term readmissions.

### Implications of all the available evidence

Patients are willing to engage both with an app targeting cardiovascular disease as well as third-party health coaches with resultant improved outpatient care transition linked to improved long-term outcomes.

Engagement was similar across demographics and socioeconomic indices. With recent digital health infrastructure expansion, similar smartphone apps may be feasible tools to optimize long-term risk reduction after hospital discharge.

