## Supplement for "Outcomes of a Smartphone-based Application with Live Health-Coaching Post-Percutaneous Coronary Intervention"

### SUPPLEMENTARY MATERIAL

#### Methods & Materials

Patients received 1 of 3 education modules (Program 1) specific to their percutaneous intervention and/or acute myocardial infarction.  All patients received the coronary artery disease (CAD) module during Program 1. Program 1 lasted approximately 30 days. Patients subsequently received additional education modules (Program 2) based on secondary patient specific conditions.  Study staff enrolled patients in one or more of the modules as applicable.  Program 2 began when subjects finished Program 1 and lasted up to 90 days depending on modules selected.

*Program 1*

| Module | Education content |
|---|---|
| CAD | • **Pathophysiology**: Heart anatomy, atherosclerosis, causes, complications<br>• **Symptoms**: angina management, cardiac precautions<br>• **Medications**: Nitroglycerin, DAPT, statins, beta blockers, ACE inhibitors<br>• **Lifestyle**: Smoking, diet, exercise, sleep, alcohol<br>• Other: Depression, intimacy, vaccines |
| Bare Metal Stent | • Warning signs, self-care, follow-up and meds specific to procedure and device |
| Drug-Eluting Stent | • Warning signs, self-care, follow-up and meds specific to procedure and device |
| Angioplasty | • Warning signs, self-care, follow-up and meds specific to procedure and device |

*Program 2*

| Module | Description of Module | Education content |
|---|---|---|
| Hyperlipidemia | *Program on managing high cholesterol based on the Therapeutic Lifestyle Changes (TLC) program* | • **Pathophysiology:** Causes, anatomy, complications<br>• **Symptoms:** CAD<br>• **Medications:** statins, aspirin<br>• **Lifestyle:** Smoking, diet, exercise, sleep, alcohol, meal planning, calories, portion control, reading labels |
| Hypertension | *Program on managing high blood pressure, including medications, diet, and other lifestyle factors.* | • **Pathophysiology:** Causes, anatomy, complications<br>• **Symptoms:** blood pressure management, cardiac precautions<br>• **Medications:** Beta blockers, ACE inhibitors, diuretics, ARBs, aspirin, statins<br>• **Lifestyle:** Smoking, diet, exercise, sleep, alcohol<br>• **Other:** Depression, intimacy, vaccines |
| Diabetes | *Program on managing diabetes, including monitoring, medications, and lifestyle factors.* | • **Pathophysiology:** Causes, anatomy, complications<br>• **Symptoms:** hypoglycemia, hyperglycemia, red flag symptoms<br>• **Medications:** Oral, insulin<br>• **Lifestyle:** Smoking, diet, exercise, sleep, alcohol, meal planning, calories, portion control<br>• **Other:** Depression, kidney disease, foot care, eye care, oral health, falls, intimacy, vaccines |
| Weight Loss | *Structured weight loss plan with a weekly curriculum* | • Calories<br>• Fiber<br>• Portion size<br>• Meal planning<br>• Goal setting<br>• Exercise<br>• Cravings<br>• Alcohol |
| Beginning Physical Activity | *Structured physical activity plan with weekly goals* | • Guidelines, safety<br>• Making a plan<br>• Injury prevention<br>• Barriers<br>• Endurance |

| | | |
|---|---|---|
| | | • Strength training<br>• Balance<br>• Flexibility |
| Smoking Cessation | *Structured smoking cessation program* | • Quit date<br>• Reasons to quit<br>• Strategies: withdrawal, cravings, meds, nicotine replacement therapy, milestones, rewards, support<br>• Weight gain<br>• Stress, depression |
| Stress | *Program on recognizing and managing stress* | • Chronic stress<br>• Stress cycle<br>• Triggers<br>• Time management<br>• Attitude, emotions<br>• Getting help<br>• Conflict negotiation<br>• Anxiety, depression |

Written patient education content was curated from evidence-based guidelines and video content delivered standardized teaching. Participants were instructed to populate their medications at enrollment and the app offered adherence tracking as well as scheduled medication reminders to promote positive health behaviors.

The health platform derived data from mobile device sensors to track activity levels, step counts and other metrics; participants could also optionally link their personal wearable devices to the app. Health advocates, who act as non-clinical coaches, were available as patient support to answer routine non-medical questions, promote engagement, support goal setting and encourage patients to reach their objectives. The health advocate acted as a co-manager to send encouraging messages and address patients' nonclinical needs to promote program retention. Any clinical concerns identified by patient inquiry, engagement patterns or survey responses were escalated by the health advocates to licensed clinicians as needed via the clinical dashboard which featured secure 2-way messaging to individuals and groups enabling efficient follow-up to answer patients' questions and outreach to encourage engagement with the care program.

Over the course of the study, participants were asked to complete survey questions on the app to assess process outcomes:

- How easy is it to navigate the Wellframe app? (0 = very hard, 10 = very easy)
- What's your favorite part about using the Wellframe app? (free text)
- What's your least favorite part about using the Wellframe app? (free text)
- What could be done to improve the Wellframe app? (free text)
- How likely are you to recommend the Wellframe app to a family member or friend? (0 = not at all, 10 = very likely)
- How successful do you think you've been in incorporating the advice from last month? (0 = not successful, 10 = is very successful)
- Using the Wellframe app has helped me feel more in control of my health. (Agree/disagree)
- Having a care manager has made me feel like I have more support in achieving my health goals. (Agree/disagree)
- How easy did you find creating an account in the Wellframe app? (0 = very hard, 10 = very easy)
- Have you had a follow-up appointment with your cardiology provider?
- After reading today's article about nutrition and CAD, what is one kind of food you want to work to eat less of in the future?
- How would you describe how much you drink alcohol? Never or very rarely; 3 drinks a month; 3 drinks a week; more than 3 drinks a day
- When you think about the changes you've made to have a healthier heart, what are you most proud of?
- Have you gone to the Emergency Room for a medical problem since you had your cath procedure?
- Have you been admitted to the hospital since you had your cath procedure?

- Have you attended an appointment with a cardiologist?
- Have you attended an appointment for cardiac rehabilitation?
- Have you gone to the Emergency Room for a medical problem since you had your cath procedure?
- Have you been admitted to the hospital since you had your cath procedure? Was this for a heart problem?
- Have you attended the first exercise session for cardiac rehabilitation?
- Have you gone to the Emergency Room for a medical problem since you had your cath procedure?
- Have you been admitted to the hospital since you had your cath procedure?

Additionally, participants received specific questions via the app survey function at discrete time points.

- At 30d: Have you attended an appointment with a cardiologist?
- At 30d: Have you scheduled an appointment for cardiac rehabilitation?
- At 60d: Have you attended an intake appointment for cardiac rehabilitation?
- At 75d: Have you attended the first exercise session for cardiac rehabilitation?

At the end of the study period, participants were called and asked about presentation to emergency rooms (including dates, reasons, subsequent admission and facility name), hospital readmission (including dates, reasons and facility names), enrollment in cardiac rehabilitation (name of facility, attendance date of intake appointment, attendance date of first exercise session), follow up with outpatient primary care provider (including what type of provider and facility location), follow up with outpatient cardiovascular clinic (including type of provider seen and facility location). Participants were also asked if they were still actively using the app and if yes, whether they would continue to use the app if given the opportunity.  Participants were subsequently instructed on deleting the app from their personal device.

**eFigure 1**: **The app included both a clinician-facing dashboard and patient-facing app.** Representative examples of clinician dashboard (center) and patient app (bottom right) using a simulated patient.

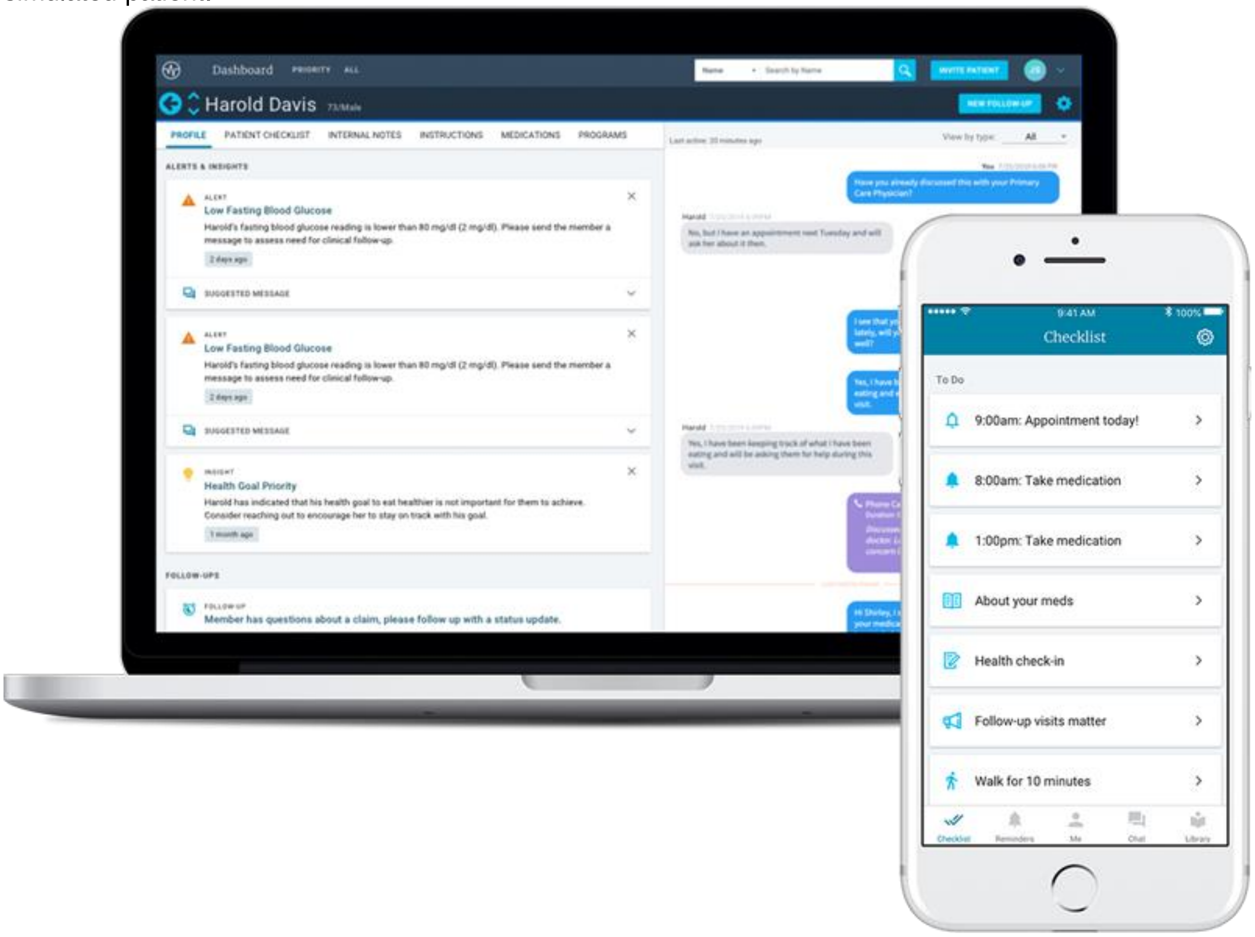

**eFigure 2**: **Exclusion criteria and process outcomes.** Process outcomes and conversion of approached patients reveals 36.4% of patients who were approached about the study enrolled. Exclusion criteria were specified prior to study enrollment start. (Missed = Not able to be seen by study staff, Elective PCI = PCI for symptomatic CAD without MI)

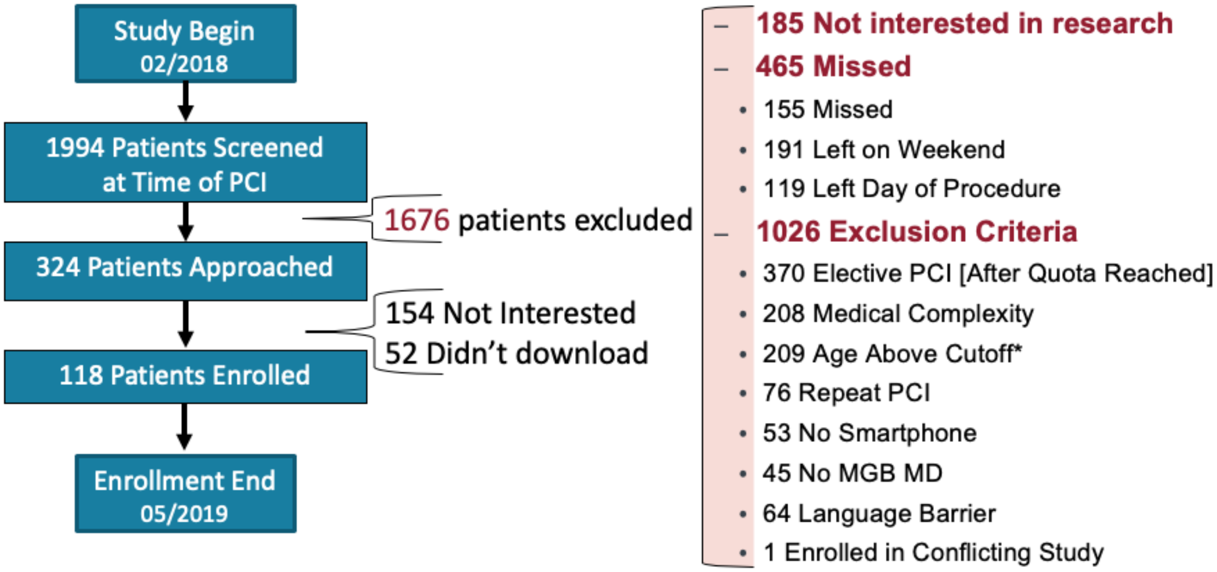


* Age cutoff was initially 70; subsequently liberalized to 85 due to patient interest.

**eFigure 3**: **Adjusted Standard Mean Differences Pre- and Post-Matching.** Covariate balance measured by standardized mean difference reflects improved balance post-matching (blue) compared to pre-matching (red). (Dotted lines represent threshold of 0.1)

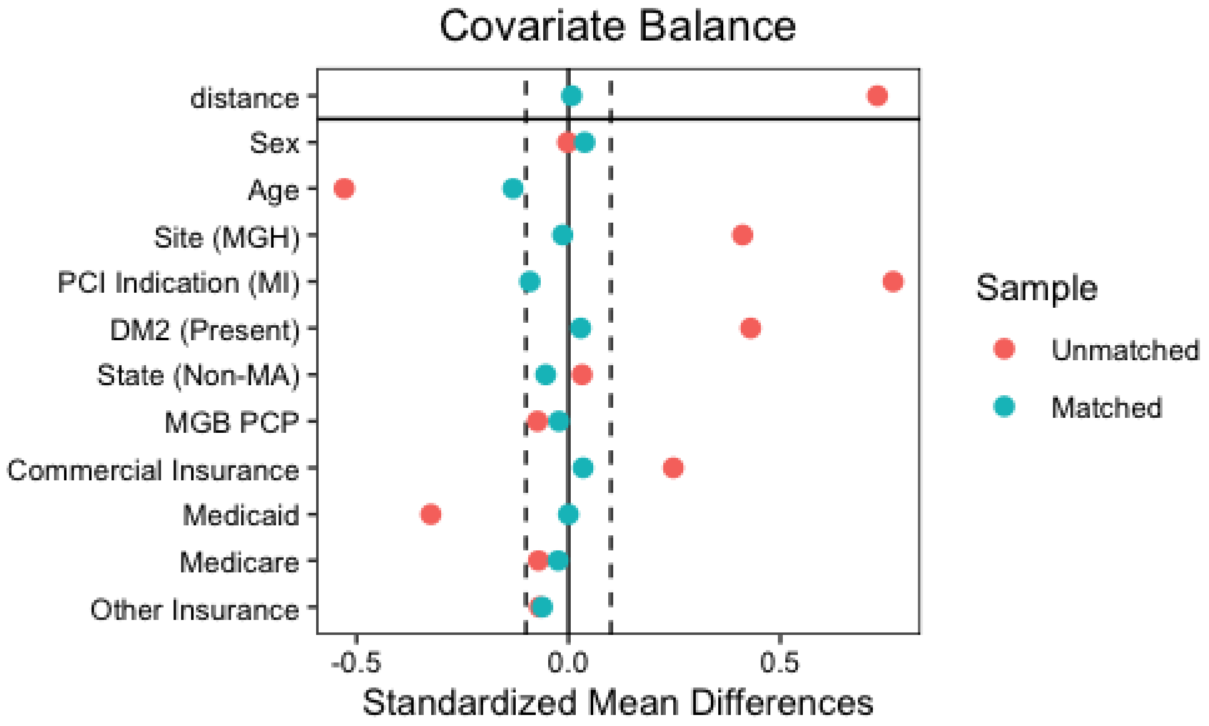

**eFigure 4**: **Histograms of patient engagement.** Engagement was parabolic with participants choosing to either engage heavily or very little. (A) In the first 30 days on the app, 30 participants engaged less than 4 days on the app whereas 39 participants engaged 28 days or more. (B) In the first 90 days on the app, 27 participants engaged less than 4 days on the app whereas 28 participants engaged 84 days or more. (Red dotted line represents 25th percentile; blue dotted line represents 75th percentile)

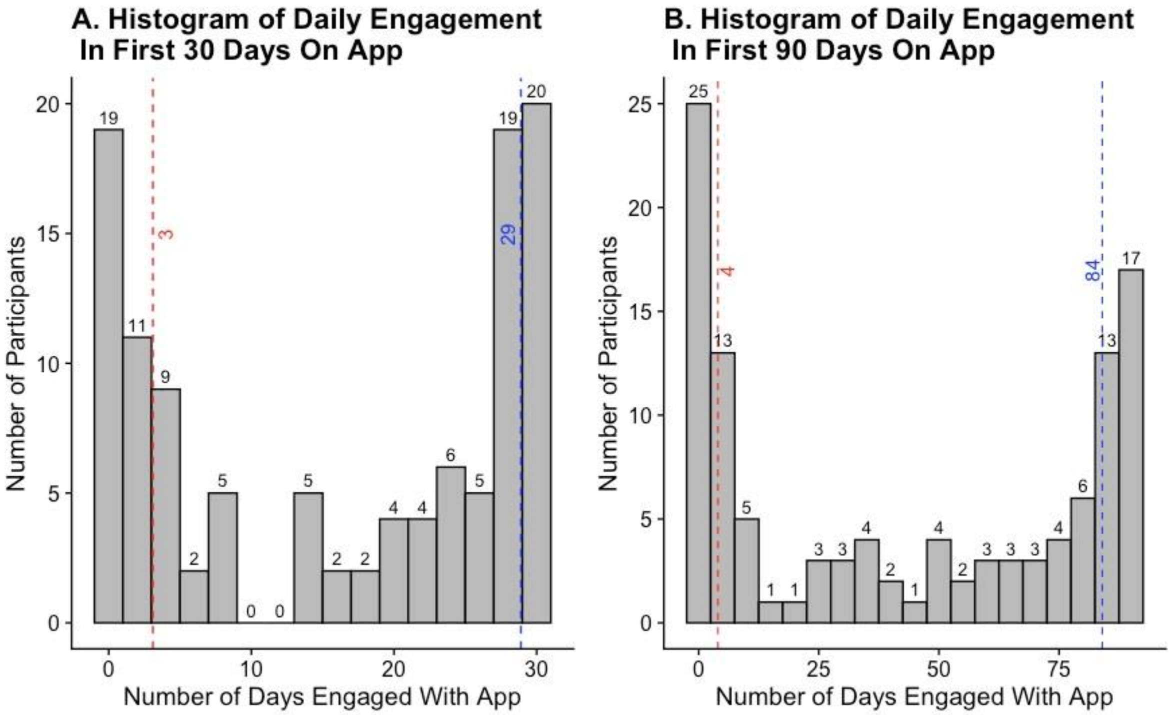

**eFigure 5**: **Within App Messaging Between Clinical Care Team and Patients.** The number of messages sent and received by patients were strongly correlated ($r^2$=0.82). (Dotted lines represent 25th, 50th and 75th percentiles, solid blue line represents linear regression, gray shading represents standard error).

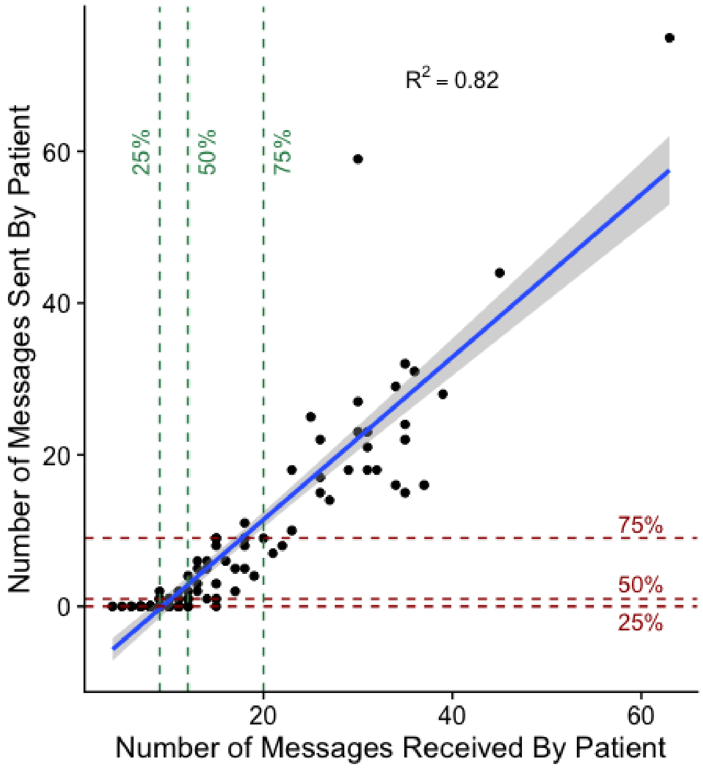

**eFigure 6**: **Engagement and traditional CV risk factors.** Correlation ($r^2$) displayed for any significant interactions (p>0.05) on Spearman correlation matrix reveals (A) high daily engagement was correlated with high weekly engagement but that daily or weekly engagement was not captured by traditional CV risk factors. And (B) content task completion was correlated with content task completion but was not captured by traditional CV risk factors.

A.

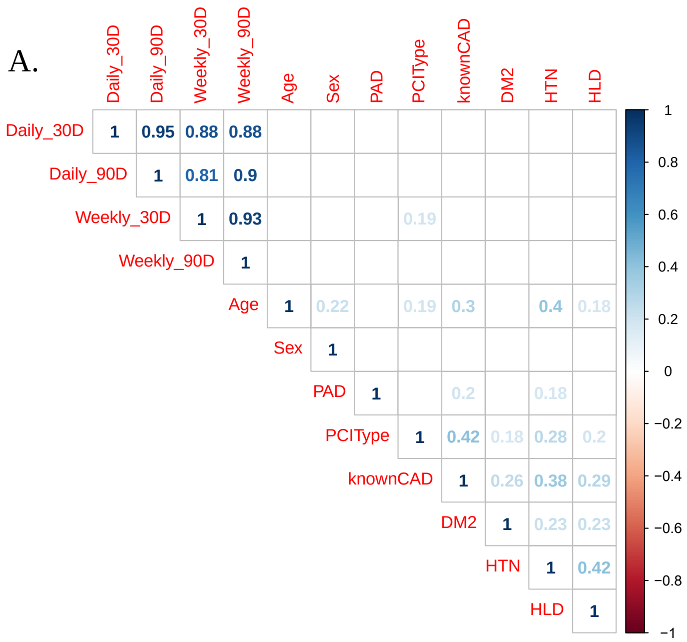


B.

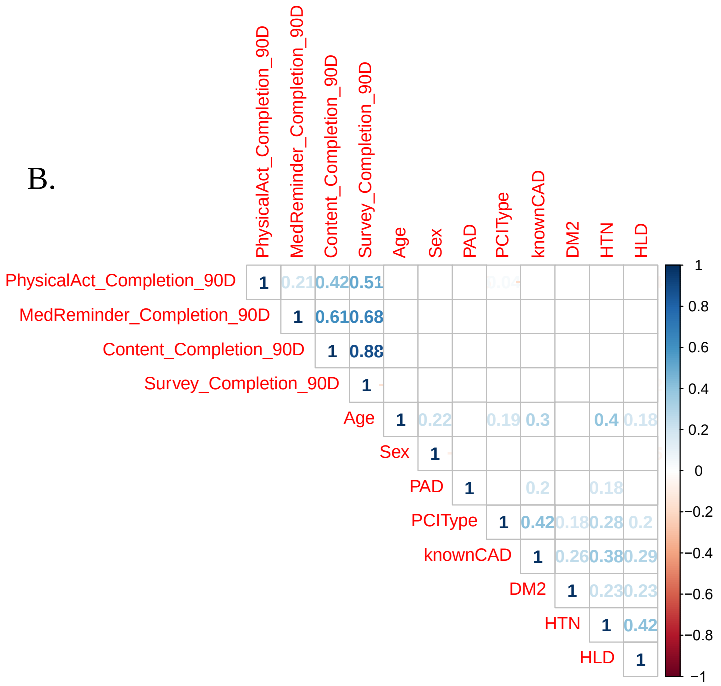

**eFigure 7: Outcomes stratified by study site.** (A) Nearly two-fold increase in attendance of cardiac rehabilitation intake at MGH site compared to historical controls from MGH and (B) slightly over two-fold increase in attendance of cardiac rehabilitation intake at BWH site compared to historical controls from BWH site. (C) Two-fold increase in 1-month outpatient cardiovascular follow up at MGH site compared to historical controls from MGH and (D) two-fold increase in 1-month outpatient cardiovascular follow up at BWH site compared to historical controls from BWH. Error bars represent confidence intervals. P-values represent Cox proportional hazards regression by site. (MGB = Mass General Brigham)

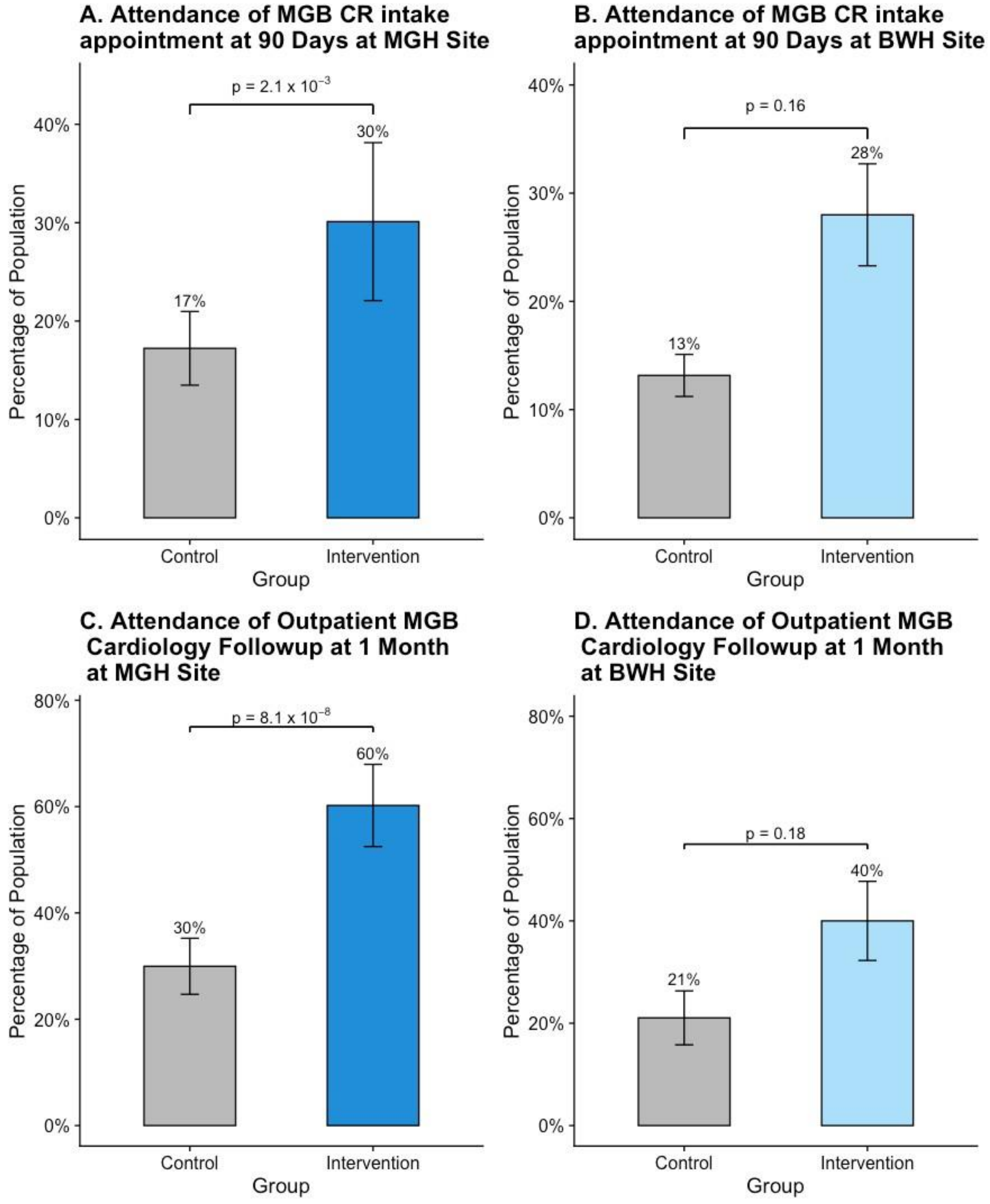

**eFigure 8**: **Daily engagement rate and clinical outcomes.** No significant differences in (A) 30- or 90-day all cause readmission, (B) 90-day cardiac rehab enrollment or (C) 1-month outpatient cardiology follow up across quartiles of engagement.

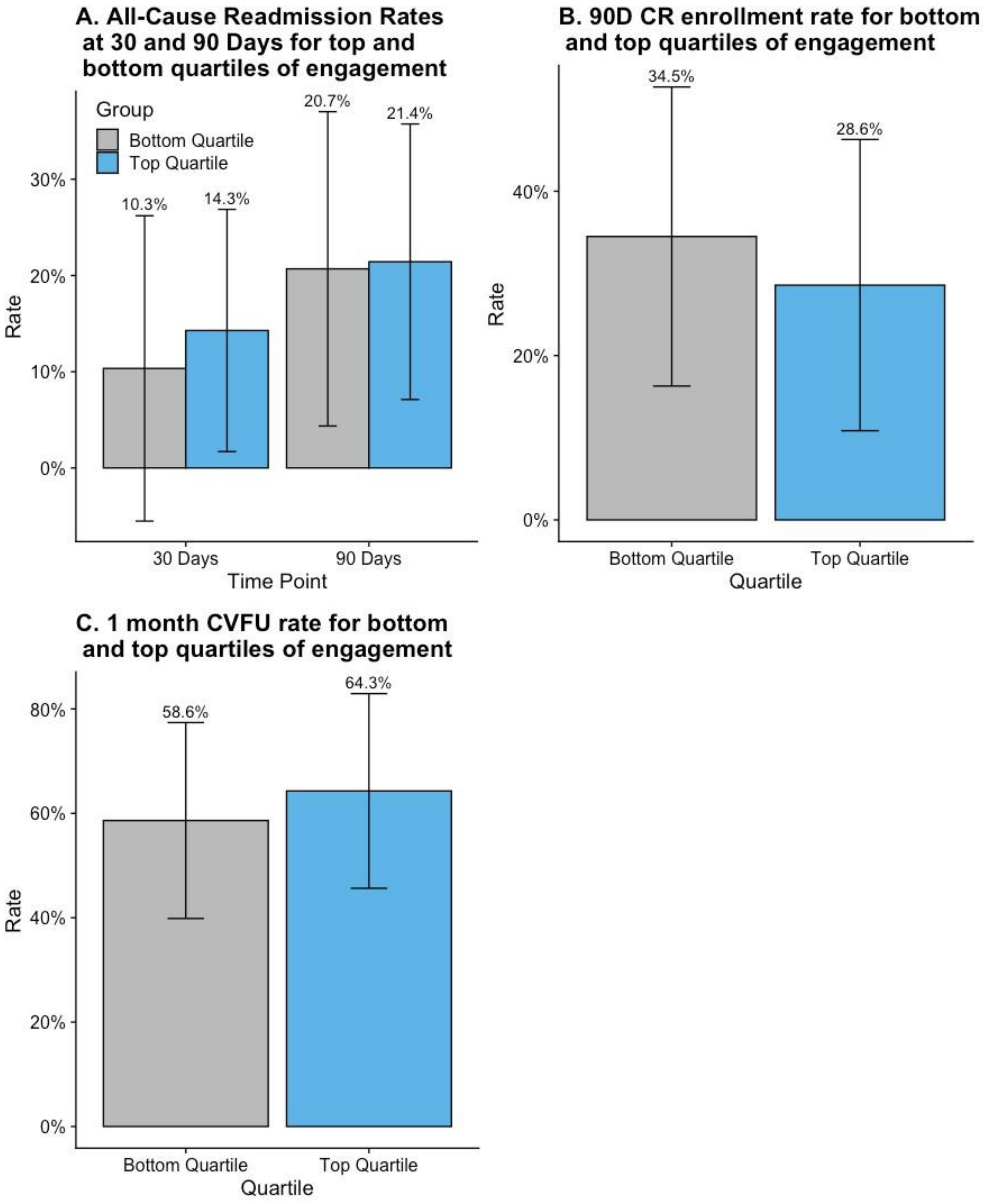

**eTable 1**: **Study metric definitions.** Descriptions of all variables involved in the study including matching criteria, outcomes criteria and engagement metrics.

| | |
|---|---|
| Age | Age at time of enrollment into study as coded into MGB EHR |
| Sex | Gender as coded into MGB EHR |
| State | State of residence (Massachusetts vs Non-Massachusetts) at time of enrollment into study as coded into MGB EHR |
| PCI Type | Reason for percutaneous coronary intervention (PCI). Myocardial infarction (MI) coded as chart diagnosis of MI or positive serologic (troponin) values prior to PCI. All other PCI without evidence of myocardial necrosis coded as elective. |
| MGB PCP | Coded "yes" for patients with PCP in MGB EMR working primarily for MGB Healthcare system. Coded "no" for patients with PCP in MGB EMR not within MGB system or missing PCP data. |
| Insurance Type | Coded based on primary insurance listed in MGB EMR. |
| Site | Coded based on location of incident PCI procedure (either Massachusetts General Hospital [MGH] or Brigham and Women's Hospital [BWH]) |
| Hypertension | Chart diagnosis of HTN prior to or at time of incident PCI |
| Hyperlipidemia | Chart diagnosis of HLD prior to or at time of incident PCI, LDL-C >129 at time of incident PCI or Triglycerides >150 at time of incident PCI |
| Type 2 Diabetes Mellitus | Chart diagnosis of DM2 prior to or at time of incident PCI, or Hemoglobin A1c > 6.4 at time of incident PCI |
| Peripheral Arterial Disease | Chart diagnosis of PAD prior to or at time of incident PCI or history of peripheral arterial intervention |
| Smoking | Chart history of smoking (codified as ever smoker (current or former smoker [quit >1 month prior to PCI]) or never smoker). |
| Known Coronary Artery Disease | Chart diagnosis of CAD, history of CABG or prior PCI prior to or at time of incident PCI |
| 30-day hospitalization | Admission date to an MGB hospital within 30 calendar days of incident PCI. |
| 90-day hospitalization | Admission date to an MGB hospital within 90 calendar days of incident PCI. |
| Cardiac reason for hospitalization | Admission for chief complaint of cardiac etiology (ie CAD, CHF, arrhythmia), admission requiring cardiac consultation or admission to a cardiology ward unit. |
| 30-day ED visit | Presentation to MGB ED or Urgent Care within 30 calendar days of incident PCI without subsequent hospitalization. |
| 90-day ED visit | Presentation to MGB ED or Urgent Care within 90 calendar days of incident PCI without subsequent hospitalization. |
| 90-day Cardiac Rehabilitation | Attendance of cardiac rehabilitation intake session at an MGB facility within 90 calendar days of incident PCI. |
| 30-day Cardiac Follow Up | Attendance of cardiac outpatient follow up appointment at an MGB facility (with MD or NP) within 30 calendar days of incident PCI |
| 30-day repeat PCI | Repeat percutaneous intervention on same or new coronary lesion within 30 days of incident PCI. |
| 90-day repeat PCI | Repeat percutaneous intervention on same or new coronary lesion within 90 days of incident PCI. |
| 90-day Stroke | Chart diagnosis of or admission for new stroke within 90 calendar days of incident PCI as noted in MGB EHR. |
| 90-day Nonfatal MI | Chart diagnosis of or admission for myocardial infarction (without subsequent death) within 90 calendar days of incident PCI as noted in MGB EHR. |
| 90-day CV Death | Documentation of death within 90 calendar days of incident PCI noted in MGB EHR with cause specified as secondary to CAD, CHF, Arrhythmia or other clear cardiovascular reason. |
| 90-day All Cause Death | Documentation of death within 90 calendar days of incident PCI noted in MGB EHR. |
| Conversion Rate | Number of patients enrolled in study out of number of total patients eligible for study. |
| Days Enrolled | Number of days between patient download of app and deletion of app by patient. |
| Medication Reminders | Number of medication reminders set up by patient (0 if no med reminders setup). |

| | |
|---|---|
| 30-day Medication Adherence | Percentage of medication reminder tasks completed within 30 days on app. (Null if no patient reminders set up) |
| 90-day Medication Adherence | Percentage of medication reminder tasks completed within 90 days on app. (Null if no patient reminders set up) |
| 30-day Content Completion | Percentage of content tasks (articles opened) completed within 30 days on app |
| 90-day Content Completion | Percentage of content tasks (articles opened) completed within 90 days on app |
| 30-day Survey Completion | Percentage of survey tasks completed within 30 days on app |
| 90-day Survey Completion | Percentage of survey tasks completed within 90 days on app |
| 30-day Weekly Patient Engagement | Percentage of days in which patient completed at least one task from the app checklist (survey, article, physical activity goal, encouragement) or sent a message to clinical team at least once over the previous 7 days within 30 days on app. |
| 90-day Weekly Patient Engagement | Percentage of days in which patient completed at least one task from the app checklist (survey, article, physical activity goal, encouragement) or sent a message to clinical team at least once over the previous 7 days within 90 days on app. |
| 30-day Daily Patient Engagement | Percentage of days in which patient completed at least one task from the app checklist (survey, article, physical activity goal, encouragement) or sent a message to clinical team within 30 days on app. |
| 90-day Daily Patient Engagement | Percentage of days in which patient completed at least one task from the app checklist (survey, article, physical activity goal, encouragement) or sent a message to clinical team within 90 days on app. |
| Patient Satisfaction | Average patient response on app satisfaction via app survey function. |
| Patient Messages Sent | Number of total messages sent by patient within 90 days on app |
| Patient Messages Received | Number of total messages sent to patient from care team within 90 days on app |
| Patient Messages Opened | Percentage of messages sent to patient that were opened |
| Patient Message Response | Percentage of messages sent to patient with at least one message response from patient |
| Patient Physical Activity | Percentage of days where patient met physical activity goal (default of 500 steps) within 90 days on app. |
| Days to First Message | Days until first message sent from care team to patient (most common) or patient to care team. |
| Clinical Messages | Percentage of patients receiving at least one message from app care team within 90 days on app. |

**Supplementary Table 2: Baseline characteristics of subjects enrolled in cardiac rehabilitation.** Demographics of participants who enrolled in cardiac rehabilitation (n=91) across both historical controls and participants enrolled in the study app were similar with the exception of insurance type (* $p < 0.05$). Fisher's exact test was used to evaluate differences across groups (†Two sample t-testing was used for continuous variables). (SD = Standard Deviation, Min = Minimum, Max = Maximum, MI = PCI for Acute Myocardial Infarction, Elective = PCI for Symptomatic CAD without Acute Myocardial Infarction, MGH = Massachusetts General Hospital, BWH = Brigham & Women's Hospital, MA = Massachusetts).

| | Historical Control Group (n=56) | Intervention Group (n=35) | p-value (Fisher's exact test) |
|---|---|---|---|
| **Age** | | | |
| Mean (SD) | 58.7 (8.27) | 62.1 (10.8) | 0.093† |
| Median [Min, Max] | 58.0 [40.0, 78.0] | 65.0 [38.0, 80.0] | |
| **Sex** | | | |
| Male | 41 (73.2%) | 27 (77.1%) | 0.806 |
| Female | 15 (26.8%) | 8 (22.9%) | |
| **PCI Type** | | | |
| MI | 41 (73.2%) | 24 (68.6%) | 0.642 |
| Elective | 15 (26.8%) | 11 (31.4%) | |
| **Site** | | | |
| MGH | 46 (82.1%) | 28 (80.0%) | 0.790 |
| BWH | 10 (17.9%) | 7 (20.0%) | |
| **Ethnicity** | | | |
| White | 43 (76.8%) | 32 (91.4%) | 0.433 |
| Black | 3 (5.4%) | 0 (0%) | |
| Hispanic | 2 (3.6%) | 0 (0%) | |
| Asian | 4 (7.1%) | 2 (5.7%) | |
| Other | 1 (1.8%) | 1 (2.9%) | |
| Unknown | 3 (5.4%) | 0 (0%) | |
| **Insurance Type** | | | |
| Commercial | 45 (80.4%) | 20 (57.1%) | 0.046* |
| Medicaid | 2 (3.6%) | 3 (8.6%) | |
| Medicare | 9 (16.1%) | 12 (34.3%) | |
| **State** | | | |
| MA | 53 (94.6%) | 34 (97.1%) | 1.000 |
| Non-MA | 3 (5.4%) | 1 (2.9%) | |
| **MGB Primary Care Provider** | | | |
| Yes | 36 (64.3%) | 27 (77.1%) | 0.246 |
| No | 20 (35.7%) | 8 (22.9%) | |

| | Historical Control Group (n=56) | Intervention Group (n=35) | p-value (Fisher's exact test) |
|---|---|---|---|
| **Diabetes Mellitus Type 2** | | | |
| Yes | 9 (16.1%) | 6 (17.1%) | 1.000 |
| No | 47 (83.9%) | 29 (82.9%) | |
| **Hypertension** | | | |
| Yes | 33 (58.9%) | 24 (68.6%) | 0.383 |
| No | 23 (41.1%) | 11 (31.4%) | |
| **Hyperlipidemia** | | | |
| Yes | 40 (71.4%) | 28 (80.0%) | 0.460 |
| No | 16 (28.6%) | 7 (20.0%) | |
| **Peripheral Arterial Disease** | | | |
| Yes | 3 (5.4%) | 0 (0%) | 0.282 |
| No | 53 (94.6%) | 35 (100%) | |
| **Known Coronary Artery Disease** | | | |
| Yes | 11 (19.6%) | 13 (37.1%) | 0.088 |
| No | 45 (80.4%) | 22 (62.9%) | |
